# How time-scale differences in asymptomatic and symptomatic transmission shape SARS-CoV-2 outbreak dynamics

**DOI:** 10.1101/2022.04.21.22274139

**Authors:** Jeremy D. Harris, Sang Woo Park, Jonathan Dushoff, Joshua S. Weitz

**Affiliations:** School of Biological Sciences, Georgia Institute of Technology, Atlanta, GA, USA; Department of Ecology and Evolutionary Biology, Princeton, NJ, USA; Department of Biology, McMaster University, Hamilton, Ontario, CA; Department of Mathematics and Statistics, McMaster University, Hamilton, Ontario, CA; M. G. DeGroote Institute for Infectious Disease Research, McMaster University, Hamilton, Ontario, CA; School of Physics, Georgia Institute of Technology, Atlanta, GA, USA; Institut de Biologie, École Normale Supérieure, Paris, France

## Abstract

Asymptomatic and symptomatic SARS-CoV-2 infections can have different characteristic time scales of transmission. These time-scale differences can shape outbreak dynamics as well as bias population-level estimates of epidemic strength, speed, and controllability. For example, prior work focusing on the initial exponential growth phase of an outbreak found that larger time scales for asymptomatic vs. symptomatic transmission can lead to under-estimates of the basic reproduction number as inferred from epidemic case data. Building upon this work, we use a series of nonlinear epidemic models to explore how differences in asymptomatic and symptomatic transmission time scales can lead to changes in the realized proportion of asymptomatic transmission throughout an epidemic. First, we find that when asymptomatic transmission time scales are longer than symptomatic transmission time scales, then the effective proportion of asymptomatic transmission increases as total incidence decreases. Moreover, these time-scale-driven impacts on epidemic dynamics are enhanced when infection status is correlated between infector and infectee pairs (e.g., due to dose-dependent impacts on symptoms). Next we apply these findings to understand the impact of time-scale differences on populations with age-dependent assortative mixing and in which the probability of having a symptomatic infection increases with age. We show that if asymptomatic generation intervals are longer than corresponding symptomatic generation intervals, then correlations between age and symptoms lead to a decrease in the age of infection during periods of epidemic decline (whether due to susceptible depletion or intervention). Altogether, these results demonstrate the need to explore the role of time-scale differences in transmission dynamics alongside behavioural changes to explain outbreak features both at early stages (e.g., in estimating the basic reproduction number) and throughout an epidemic (e.g., in connecting shifts in the age of infection to periods of changing incidence).

## 1 Introduction

The role of asymptomatic carriers in driving epidemic dynamics has remained a key question throughout the SARS-CoV-2 pandemic [2, 6, 15, 23]. Asymptomatic carriers have reduced the effectiveness of non-pharmaceutical interventions [4, 7, 12, 15, 16, 35], and have made it more difficult to obtain unbiased estimates of disease severity, including infection fatality ratios [28]. Although several studies have estimated the prevalence of asymptomatic SARS-CoV-2 infections in various settings [18–22], there is still considerable uncertainty in how the transmission dynamics of asymptomatic individuals differ from those of symptomatic individuals. Modeling studies have typically assumed that asymptomatic and symptomatic individuals are infected for an equal amount of time. Some studies have further accounted for the possibility that asymptomatic individuals may transmit less than symptomatic individuals, but the range of assumptions vary widely (from 90% less transmissible [11] to equally transmissible [17]).

Prior work has shown that individual-level differences in transmission time scales between asymptomatic and symptomatic transmission can have important implications for estimates of epidemic dynamics during the exponential growth phase [25]. For example, if asymptomatic individuals are able to transmit for a longer period of time than symptomatic individuals, the proportion of new infections attributable to asymptomatic transmission will be lower than predicted based on their intrinsic infectiousness because shorter transmission intervals drive the spread during the epidemic growth phase. Under the same scenario, failing to account for differences in time scales of asymptomatic and symptomatic transmission can lead to underestimation of the basic reproduction number (i.e., the average number of secondary infections caused by a primary case [1, 9, 10, 31]) from the epidemic growth rate [25]. These differences in transmission time scales can be driven by both biological (e.g., longer viral shedding period [19]) and behavioral factors (e.g., self-isolation of symptomatic individuals).

The impact of transmission time-scale differences on epidemic inference can be approached using a generation interval-based framework [13]. The generation interval, defined as the time between when an individual is infected and when that individual transmits to another person, connects individual-level transmission time scales with population-level measures of disease spread. [5, 30, 32, 33]. For example, given an observed epidemic growth rate, a disease with longer generation intervals on average will be associated with a higher reproduction number [24]. This result applies here: if asymptomatic cases have longer generation intervals the overall average generation interval is increased [25]).

Symptomaticity of infections may also correlate with transmission outcomes. That is, new infections caused by asymptomatic transmission may be more likely to remain asymptomatic than new infections caused by symptomatic individuals, leading to correlations between disease statuses of the infector and of the infectee. Such correlations might arise from dose-dependent responses: recent animal-model studies have shown such responses to COVID-19 infection, with higher initial viral inoculum associated with both increased viral shedding and more severe outcomes [14, 29]; data from animal model studies of other human coronaviruses, including SARS-CoV, have shown similar trends [34]. If symptomatic infections typically shed more infectious virus than asymptomatic infections, then the initial viral dose from symptomatic transmission would be higher on average than from asymptomatic transmission, making symptomatic infections more likely, and generating correlations between disease statuses of the infector and the infectee.

Correlations might also arise from age-dependent assortativity in mixing patterns and variation in symptomaticity. Higher contact rates among individuals of similar ages (e.g., in schools) cause more transmission within similar age groups (as opposed to between different age groups). As disease symptomaticity of SARS-CoV-2 varies with age [8, 35], disease statuses of infectees may effectively correlate with disease statuses of their infectors. Irrespective of the mechanism, we predict that such correlations can have important dynamical consequences when they are coupled with the effects of differences in time scales of transmission: if the proportion of new infections attributable to asymptomatic transmission changes over the course of an epidemic, these correlations may amplify changes in the realized proportion of asymptomatic incidence.

In this study, we examine the impacts of individual-level differences between asymptomatic and symptomatic transmission on population-level disease dynamics. First, we consider the possibility that asymptomatic individuals may transmit more slowly (i.e., have longer generation intervals on average) than symptomatic individuals. We show that such slow transmission by asymptomatic individuals would increase the realized proportion of asymptomatic transmission during periods when total transmission is declining—a robust pattern whether the decline is driven by susceptible depletion or by changes in effective contacts. Second, we account for the correlations between transmission outcomes and individual disease statuses. In this case, we find that the proportion of asymptomatic transmission as well as a new effect: the proportion of asymptomatic *incidence* can also increase as the epidemic declines.

Finally, we study the dynamics of an age-dependent model that includes assortative mixing and a greater proportion of asymptomatic infections in younger than older individuals. In this example, the average age of an incident infection increases as the epidemic progresses, because of faster depletion of susceptibles in younger age classes. Because the probability of symptomatic infections increases with age, this leads to changes in the realized proportion of asymptomatic transmission and incidence. In an intervention scenario, the average age of an incident infection remains nearly constant when asymptomatic and symptomatic generation-interval distributions are identical. However, when asymptomatic generation intervals are longer, the average age of an incident infection decreases as the epidemic decays, causing the realized proportion of asymptomatic incidence to increase. Together, these analyses demonstrate the potential for individual-level variation in transmission dynamics to shape the realized proportion of asymptomatic transmission and incidence throughout an epidemic.

## 2 Methods

### 2.1 SEIR model of asymptomatic transmission with a fixed intrinsic proportion of asymptomatic incidence

We study the impact of differences in asymptomatic and symptomatic generation-interval distributions on epidemic dynamics using a series of Susceptible-Exposed-Infectious-Recovered (SEIR) models. Once infected, susceptible individuals enter an exposed but latent stage, during which they cannot transmit. The first model assumes that a fixed proportion *p*—which we refer to as the intrinsic proportion of asymptomatic infections—of newly infected individuals remains asymptomatic over the course of infection while transmitting at rate *β*_*a*_. The remaining proportion 1 *− p* develops symptoms after the exposed period and transmit at rate *β*_*s*_. Then, the proportion of individuals in each compartment can be described by the following set of equations:

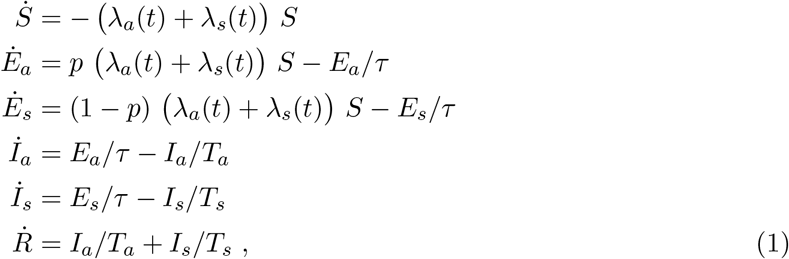

where subscripts denote asymptomatic (*a*) vs. symptomatic (*s*) classes. Here,

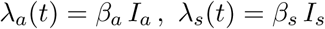

denote the forces of infection caused by asymptomatic and symptomatic individuals, respectively, 1*/τ* is the mean exposed period, and *T*_*a*_ (*T*_*s*_) is the mean duration of asymptomatic (symptomatic) infectious periods. Since infectious period is assumed to be exponentially distributed, the mean generation interval is equal to the sum of the mean exposed and infectious periods [27, 30, 32].

For this model, the subgroup reproduction number of asymptomatic (symptomatic) individuals is given by *ℛ*_0,*a*_ = *β*_*a*_ *T*_*a*_ (*ℛ*_0,*s*_ = *β*_*s*_ *T*_*s*_) and defined as the number of secondary infections caused by a single asymptomatically (symptomatically) infected individual in a fully susceptible population. The basic reproduction number of the system is the weighted average of the two subgroup reproduction numbers:

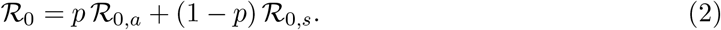

Then, we can define the intrinsic proportion of asymptomatic transmission, which represents the relative contribution of asymptomatic transmission towards the basic reproduction number [25]:

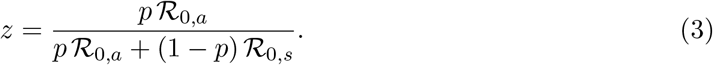

We also define the realized proportion of asymptomatic transmission, *q*(*t*), the proportion of new infections caused by asymptomatically infected individuals at time *t*:

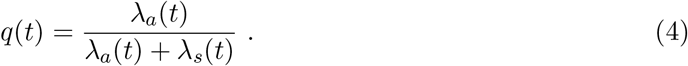

If asymptomatic and symptomatic individuals have identical generation-interval distributions, then *q*(*t*) = *z*. However, if the generation-interval distributions differ, then the realized proportion of asymptomatic transmission can systematically differ from the intrinsic proportion of asymptomatic transmission, not only during exponential growth but also as the epidemic progresses.

Here, we generalize prior work [25], which focused on the initial exponential growth phase, and we explore how the realized proportion of asymptomatic transmission changes over the course of an epidemic when we assume that asymptomatic infections have longer infectious periods (and therefore longer generation intervals). To do so, we assume the mean exposed period is *τ* = 3 days for both groups and fix the mean infectious period to *T*_*s*_ = 5 days for symptomatic infections. We then explore changes in the mean infectious period between *T*_*a*_ = 5 to *T*_*a*_ = 8 days for asymptomatic infections. To compare across simulations, we fix the intrinsic proportion of asymptomatic infections (*p* = 0.4). We set the subgroup reproduction numbers are equal, i.e., *ℛ*_0,*a*_ = *ℛ*_0,*s*_, and vary transmission such that the exponential growth rate is matched across simulations (*r* = 0.14*/*day). We also provide a supplemental figure that shows simulations when transmission rates are equal. We show simulations for both susceptible depletion and intervention scenarios. For intervention, rates of asymptomatic and symptomatic transmission are reduced by an equal factor such that the effective reproduction number is reduced over a period of 30 days starting from 70 days into the simulation. The mitigation intensities are chosen so that the final effective reproduction numbers match those observed in the susceptible depletion scenario for each infectious period. See Table 1 for parameter descriptions and Figure S1 for model schematic.

**Table 1.**
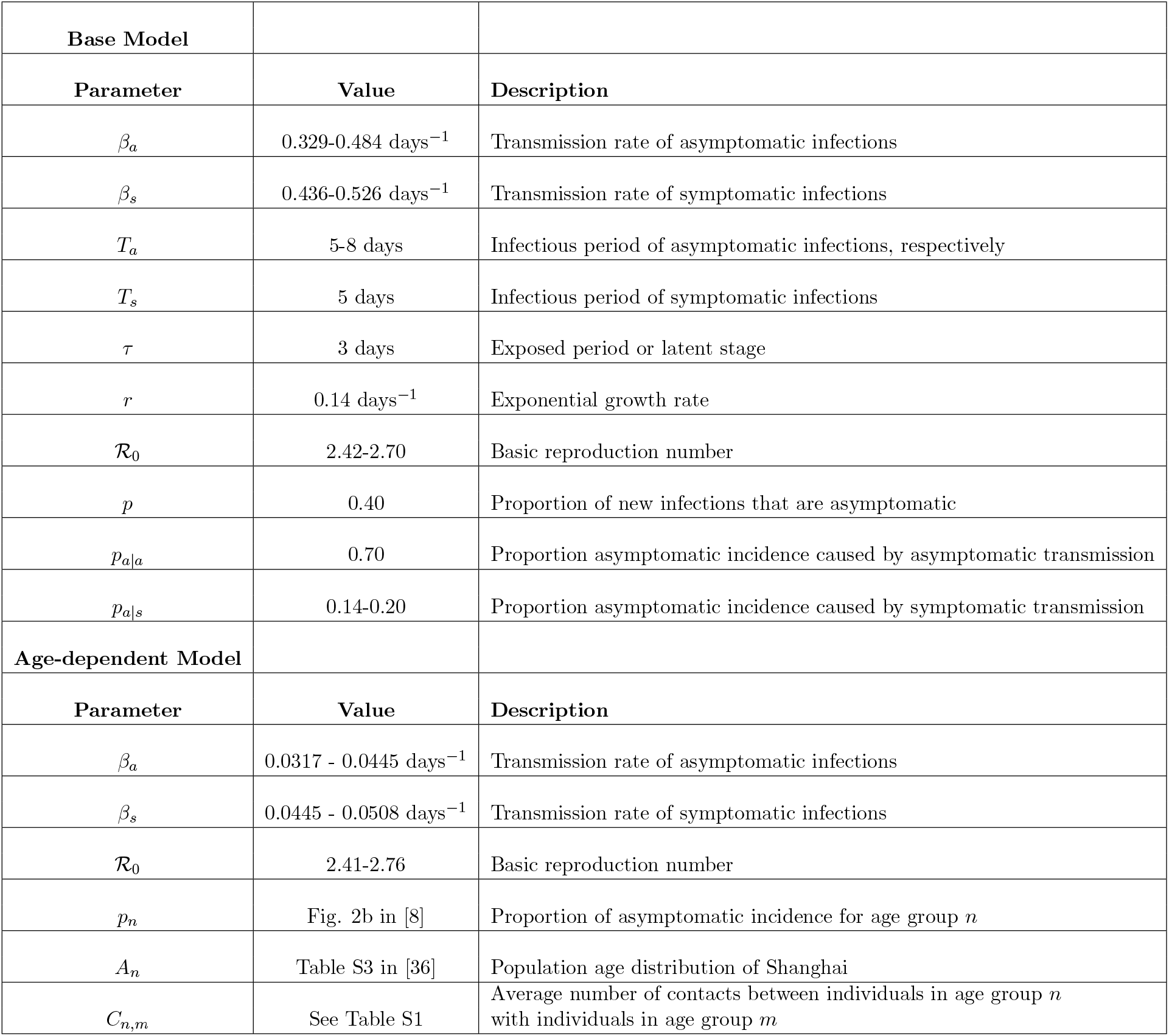
Parameters, Values, Descriptions. SEIR models with asymptomatic and symptomatic infections. Parameters for base models and age-dependent model.

### 2.2 SEIR model of asymptomatic transmission with correlations between transmission outcomes and disease statuses

Next, we introduce correlations between transmission outcomes and disease statuses: that is, transmission from asymptomatic (symptomatic) individuals is more likely to lead to new, asymptomatic (symptomatic) infections. To study the effects of such correlations on epidemic dynamics, we extend the model in Eq. (1):

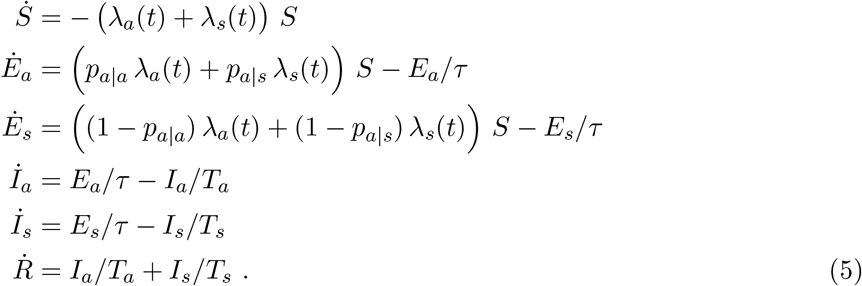

Here, *p*_*a*|*a*_ (*p*_*a*|*s*_) is the proportion of new asymptomatic infections caused by asymptomatic (symptomatic) transmission, whereas *p*_*s*|*a*_ = 1 *− p*_*a*|*a*_ (*p*_*s*|*s*_ = 1 *− p*_*a*|*s*_) is the proportion of new symptomatic infections caused by asymptomatic (symptomatic) transmission. When *p*_*a*|*a*_ = *p*_*a*|*s*_, the model in Eq. (5) reduces to the model in Eq. (1).

The realized proportion of asymptomatic transmission is still given by Eq. (4). The realized proportion of asymptomatic *incidence* needs to account for the new correlations:

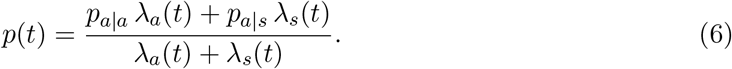

We explore the effect of correlations between transmission outcomes and disease statuses by letting *p*_*a*|*a*_ *> p*_*a*|*s*_ while holding the initial realized proportion of asymptomatic incidence fixed at *p* = 0.4, and the exponential growth rate fixed at *r* = 0.14*/*day across simulations. As before, we assume the subgroup basic reproduction numbers are equal, *ℛ*_0,*a*_ = *ℛ*_0,*s*_, and provide a supplemental figure that shows simulations when transmission rates are equal. See Table 1 for parameter descriptions and Figure S1 for model schematic.

### 2.3 SEIR model of asymptomatic transmission with assortative mixing and variation in the chance of symptomatic infection by age

Finally, we study an age-stratified model as an example of how correlations might arise between transmission outcomes and disease statuses. The model couples age-dependent assortative mixing patterns with variation in the proportion of asymptomatic (vs. symptomatic) outcomes that increase with age. Hence, if younger individuals are more likely to remain asymptomatic and assortatively mix with younger individuals, then asymptomatic infections will effectively cause more asymptomatic infections than symptomatic infections. To examine these potential age-dependent effects, we stratify the population into age groups spanning intervals of 10 years, going from 0-9 (*n* = 1) up to 60-69 (*n* = 7) with the last group being 70+ (*n* = 8). Each age group *n* consists of 6 compartments (*S*_*n*_, *E*_*n,a*_, *E*_*n,s*_, *I*_*n,a*_, *I*_*n,s*_, and *R*_*n*_) representing the number of individuals in each disease state such that *S*_*n*_ + *E*_*n,a*_ + *E*_*n,s*_ + *I*_*n,a*_ + *I*_*n,s*_ + *R*_*n*_ = *A*_*n*_, where *A*_*n*_ is the population size in age group *n*.

We model the contact patterns between the age groups by including an empirically estimated contact matrix, 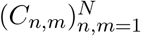, where *C*_*n,m*_ is the average number of contacts (per day) that individuals in age group *n* make with individuals in age group *m* [36]. In our simulations, we use the baseline contact estimates of Shanghai that were empirically estimated prior to the COVID-19 outbreak, which shows a high degree of age-dependent assortative mixing [36] (See Table S1). To be consistent with estimates of contact rates, we let *A*_*n*_ be the age distribution of the population of Shanghai. We vary the proportion of asymptomatic incidence with respect to age, *p*_*n*_ [8]. In this model, we introduce intervention earlier than in the previous models (50 days into the simulation) to ensure limited susceptible depletion across all of the age groups. We match the exponential growth rate across simulations (*r* = 0.14*/*day), and assume that *ℛ*_0,*a*_ = *ℛ*_0,*s*_. See Table 1 for parameter descriptions.

Then the number of individuals in each age group and disease state can be described by the following set of equations:

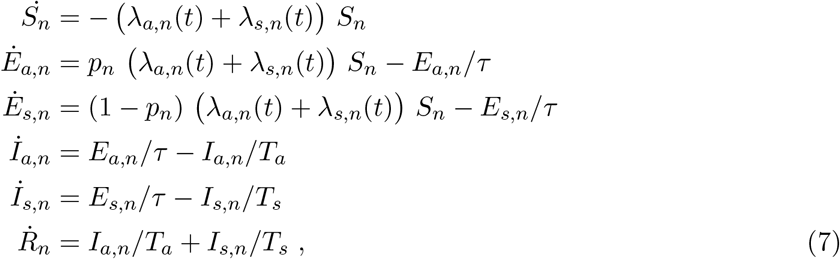

where the forces of infection for each age group *n* due to asymptomatic (*a*) and symptomatic (*s*) transmission are given by

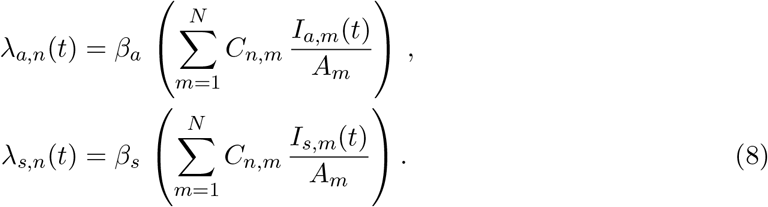

As before, we compute the realized proportion of asymptomatic transmission over time

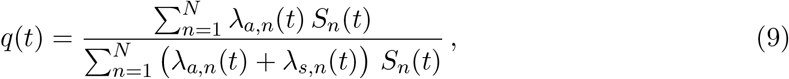

as well as the realized proportion of asymptomatic incidence over time,

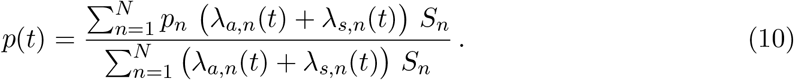

We also calculate the average age of an incident infection over time:

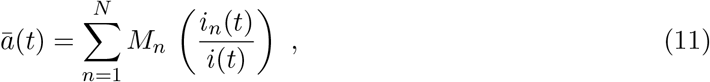

where *M*_*n*_ is the midpoint of age group *n, i*_*n*_(*t*) = (*λ*_*a,n*_(*t*) + *λ*_*s,n*_(*t*)) *S*_*n*_ is the incidence of age group *n*, and 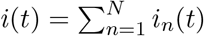 is the total incidence across all age groups.

## 3 Results

### 3.1 Effects of differences asymptomatic and symptomatic generation intervals

We first study the effects of differences asymptomatic and symptomatic generation intervals using the SEIR model by fixing intrinsic proportion of asymptomatic incidence, *p*. To do so, we simulate the model under two scenarios: (1) ‘Susceptible Depletion’, where the epidemic spreads without mitigation, and (2) ‘Intervention’, where intrinsic transmission rates of asymptomatic and symptomatic infections are exogenously reduced by the same proportion Since the exponential growth rate is matched across simulations, the incidence curves start off identically across all simulations (Figure 1A,E). When the mean infectious period of asymptomatic individuals is longer than that of symptomatic individuals, the incidence curves decay more slowly (*T*_*a*_ = 6 days, purple and *T*_*a*_ = 8 days, light blue). In this case the realized proportion of asymptomatic transmission, *q*(*t*), also increases over time because slower generation intervals of asymptomatic individuals become relatively more important during the decay phase (Figure 1B,F). The differences in the mean infectious periods of asymptomatic and symptomatic individuals do not affect the realized proportion of asymptomatic incidence (Figure 1C,G). Since changes in the realized proportion of asymptomatic transmission, *q*(*t*), are determined by the epidemic growth/decay rate [25], we are able to match the magnitude of changes in the realized proportion of asymptomatic transmission between susceptible depletion and intervention scenarios by matching their final effective reproduction numbers (Figure 1B,F).

**Figure 1.**
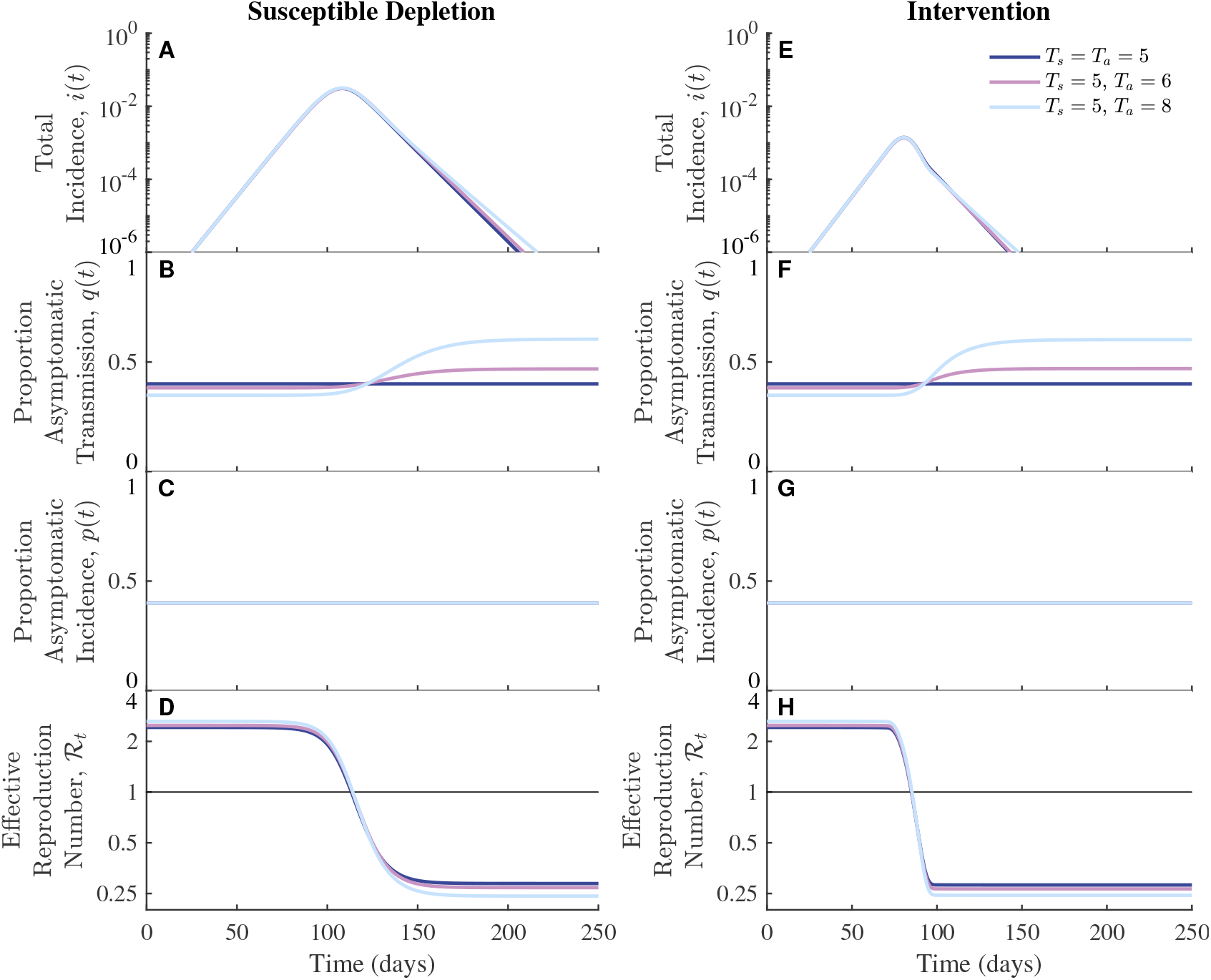
Effects of differences in asymptomatic and symptomatic generation-interval distributions on the population-level dynamics of asymptomatic infections. We fix the infectious period of symptomatic infections, *T*_*s*_ = 5 days and increase the infectious period of asymptomatic infections from *T*_*a*_ = 5 days (dark blue), *T*_*a*_ = 6 days (purple), *T*_*a*_ = 8 days (light blue). (A-D) Without intervention the epidemic spreads through the susceptible population unhindered. As total incidence decreases, the proportion of asymptomatic transmission increases over time when asymptomatic infectious periods are longer than symptomatic infectious periods (purple and light blue). (E-H) With intervention, the reproduction number is reduced over a period of 30 days with mitigation intensities such that the final effective reproduction numbers match those in the susceptible depletion case. Across all simulations, the intrinsic proportion of asymptomatic incidence is *p* = 0.40, and the exponential growth rate is *r* = 0.14*/*day (Methods). Other parameter values: *ℛ*_0_ = 2.42, *β*_*a*_ = *β*_*s*_ = 0.484 days^*−*1^ (dark blue); *ℛ*_0_ = 2.48, *β*_*a*_ = 0.416 days^*−*1^, *β*_*s*_ = 0.497 days^*−*1^ (purple); *ℛ*_0_ = 2.63, *β*_*a*_ = 0.329 days^*−*1^, *β*_*s*_ = 0.526 days^*−*1^ (light blue).

We further investigate how the magnitude and timing of intervention affect the realized proportion of asymptomatic transmission. When we fix the final effective reproduction and vary the mitigation onset time, the resulting realized proportions of asymptomatic transmission *q*(*t*) show similar trajectories and identical asymptotic values across all scenarios because *q*(*t*) is determined by the exponential decay rate (Figure S2A-D). When we fix the mitigation onset time and vary the final effective reproduction number, more intense interventions cause incidence to decay faster, which in turn corresponds to a larger increase in the realized proportion of asymptomatic transmission (Figure S2E–G).

We obtain similar results when we assume the transmission rates are equal for asymptomatic and symptomatic infections (Figure S3), rather than assuming the reproduction numbers are equal (as in Figure 1). The magnitude of changes in the proportion of asymptomatic transmission are similar across both cases (equal transmission rates vs. equal basic reproduction numbers). The one difference is in the initial realized proportion of asymptomatic transmission. When the reproduction numbers are equal, the initial realized proportion of asymptomatic transmission is lower than the intrinsic proportion of asymptomatic incidence (Figure 1B,F), whereas when transmission rates are equal, the initial realized proportion of asymptomatic transmission is greater than the intrinsic proportion of asymptomatic incidence (Figure S3B,F).

### 3.2 Effects of correlations between transmission outcomes and disease statuses

Next, we study the effects of correlations between transmission outcomes and disease statuses of the infectors on epidemic dynamics across two scenarios (with and without intervention). When the mean infectious periods of asymptomatic and symptomatic infections are equal, correlations between transmission outcomes and disease statuses have no effect on epidemic dynamics: the incidence curves (Figure 2A,E; dashed dark blue) are identical to those in the case with fixed intrinsic proportion asymptomatic incidence (indicated by ‘*p* = 0.40’). The realized proportions of asymptomatic transmission and incidence also remain constant over time—in this case, both proportions are equal to the intrinsic proportion asymptomatic incidence (*p* = 0.40) because asymptomatic and symptomatic individuals have identical reproduction numbers (Figure 2B,F and C,G; dashed dark blue). When asymptomatic individuals have longer infectious periods (and therefore longer generation intervals), correlations between transmission outcomes and disease statuses exaggerate the effect of differences in the transmission time scale—incidence of infection decays more slowly (Figure 2A,E; light blue curves) and the realized proportion of asymptomatic transmission increases by a greater amount (Figure 2B,F; light blue curves). In particular, an increase in asymptomatic transmission also causes the proportion of asymptomatic incidence to increase because transmission from asymptomatically infected individuals are more likely to result in new asymptomatic infections (Figure 2C,G; dashed light blue).

**Figure 2.**
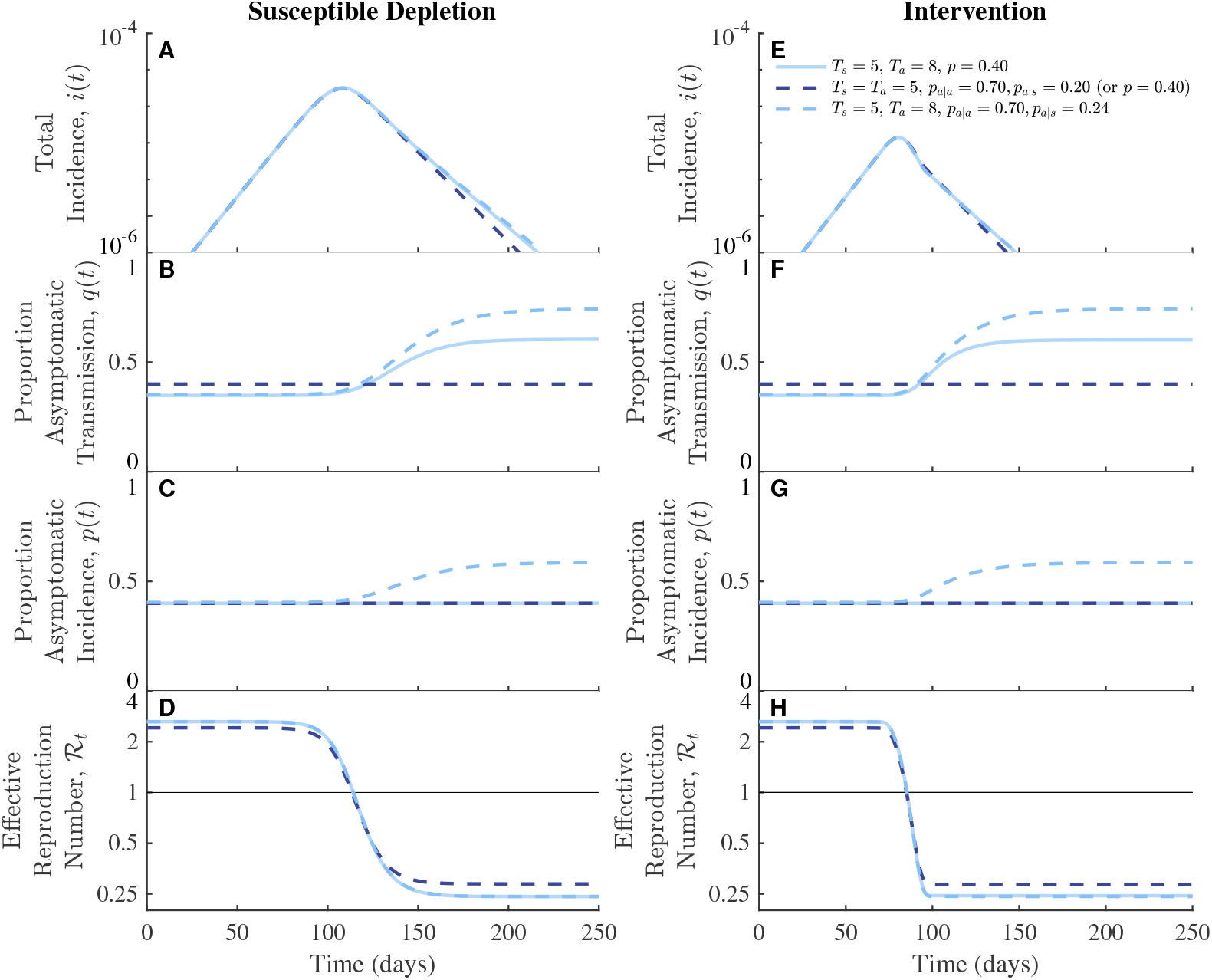
Effects of transmission correlations and generation-interval differences on the population-level dynamics of asymptomatic infections. We fix the infectious period of symptomatic infections to *T*_*s*_ = 5 days and increase the correlation between transmission and disease status. For comparison, we include *T*_*a*_ = 8 days with fixed intrinsic proportion asymptomatic incidence (*p* = 0.40; solid light blue) which is the same as in Figure 1. When generation intervals are equal, dynamics are identical for the correlated and uncorrelated case (dashed dark blue) if the overall transmission proportion is matched. In both the susceptible depletion (A-D) and intervention cases (E-H), longer generation intervals of asymptomatic transmission (*T*_*a*_ = 8 days) lead to increases in the realized proportion of asymptomatic transmission over time (B,F, light blue curves). Coupling correlations between transmission and disease statuses with longer generation intervals of asymptomatic transmission cause the realized proportion of asymptomatic incidence to increase over time (C,G, dashed light blue). The intrinsic proportion of asymptomatic incidence, *p* = 0.4 and the exponential growth rate, *r* = 0.14 days^*−*1^, are matched across all simulations (Methods). Parameter values: *p* = 0.40 (solid light blue), *p*_*a*|*a*_ = 0.70, *p*_*a*|*s*_ = 0.20 (dashed dark blue); *p*_*a*|*a*_ = 0.70, *p*_*a*|*s*_ = 0.14 (dashed light blue). Other parameter values are the same as in Figure 1 for corresponding colors.

### 3.3 Effects of age-dependent mixing

Finally, we consider the effects of coupling age-dependent assortativity and symptomaticity as an example of how correlations might arise between transmission outcomes and disease statuses. Since symptomaticity is correlated with age, and since individuals are more likely to mix with other individuals of similar age groups, relatively higher proportions of asymptomatic (symptomatic) secondary infections are due to transmission from asymptomatic (symptomatic) primary infections. To investigate the effect of age-dependent heterogeneity on the dynamics of asymptomatic infections, we parametrise an age-dependent SEIR model by allowing the contact rates and the proportions of asymptomatic incidence to vary across age groups (Methods).

First, we consider age-dependent assortative mixing in contacts and examine the effects of introducing age-dependent variation in proportion of asymptomatic infection (Figure S5). In the absence of intervention, the average age of an incident infection increases as the epidemic progresses in this example because higher contact rates of younger individuals drive faster susceptible depletion (Figure S5B). An increase in the mean age of infection translates to a decrease in both the proportion of asymptomatic incidence (Figure S5C) and transmission (Figure S5D). In contrast, intervention prevents significant susceptible depletion of each age group, and thus, the age distribution of incident infections, and therefore proportions of asymptomatic transmission and infections, remains roughly constant over time (Figure S5G–I).

When we increase the mean infectious period of asymptomatic individuals, longer generation intervals from young, asymptomatic individuals become relatively more important during the decay phase; therefore, the mean age of an incident infection decreases (Figure S5B, G; dashed light blue) and the proportions of asymptomatic transmission and incidence increase (Figure S5C, D, H, I; dashed light blue).

**Figure 3.**
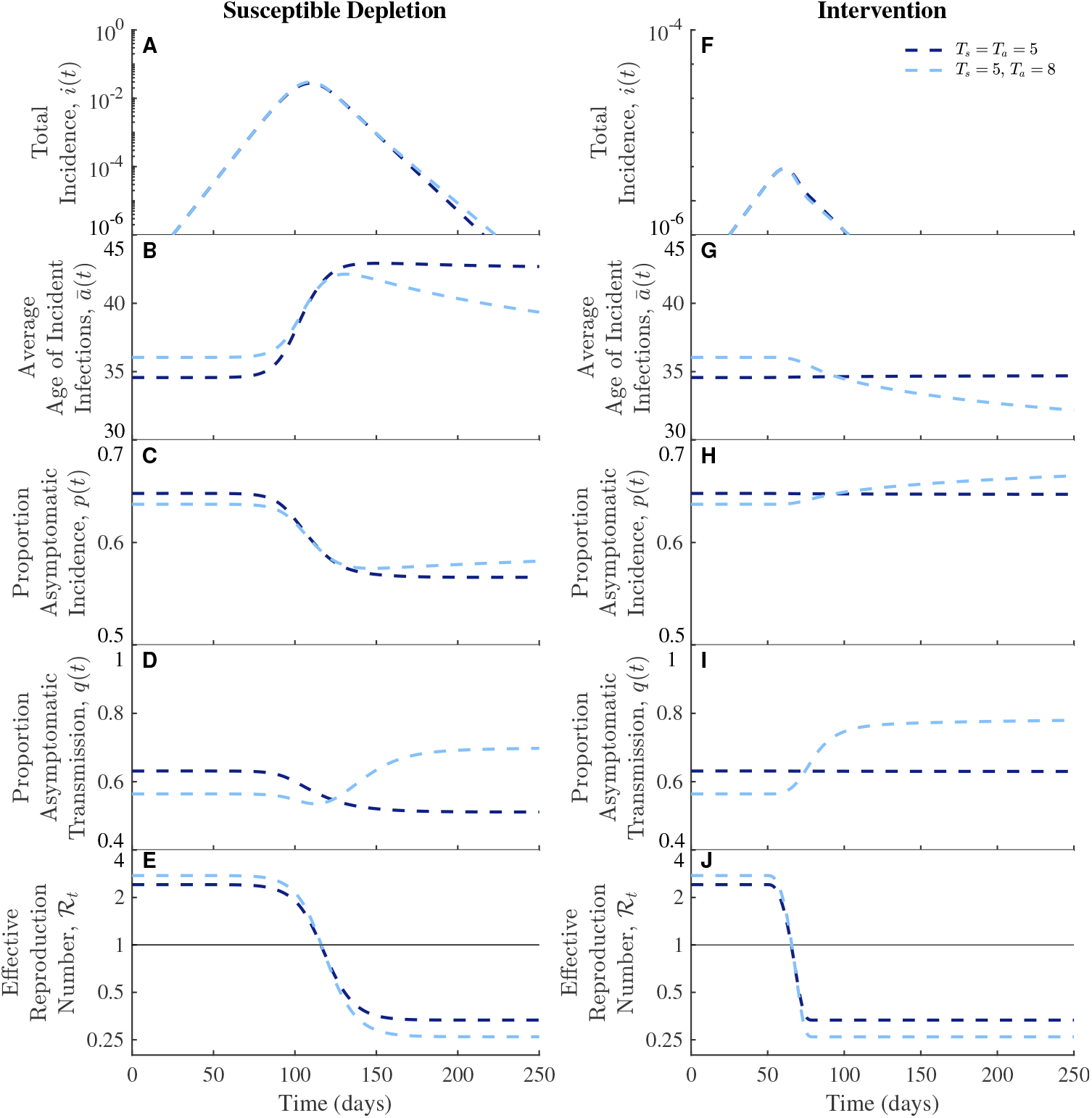
Effects of age-dependent mixing and generation-interval differences on the population-level dynamics of asymptomatic infections. We fix the symptomatic infectious period to *T*_*s*_ = 5 days and compare when the asymptomatic infectious period is equal to (*T*_*a*_ = *T*_*s*_ = 5 days) or longer than the symptomatic infectious period (*T*_*a*_ = 8 days). We show susceptible depletion (A-E) and intervention scenarios (F-J). As the average age of an incident infection changes over time (C,H), so do the realized proportions of asymptomatic incidence (C,H) and transmission (D,I). Across all simulations, the intrinsic proportion of asymptomatic incidence is 0.648, and the exponential growth rate is *r* = 0.14 days^*−*1^ (Methods). Other parameter values: *ℛ*_0_ = 2.41, *β*_*a*_ = *β*_*s*_ = 0.0445 days^*−*1^ (dashed dark blue); *ℛ*_0_ = 2.76, *β*_*a*_ = 0.317 days^*−*1^, *β*_*s*_ = 0.0508 days^*−*1^ (dashed light blue).

## 4 Discussion

Using a series of nonlinear epidemice models, we found that time-scale differences in transmission dynamics between symptomatic and asymptomatic individuals can shape population-level epidemic dynamics. In particular, when asymptomatic individuals transmit for longer, the proportion of asymptomatic transmission tends to increase as the epidemic decays because longer generation intervals of asymptomatic transmission become more important; this result generalizes earlier work, which illustrated the same effect for the initial exponential growth phase [25]. Further accounting for the possibility that asymptomatic individuals are more likely to generate asymptomatic infections can amplify this effect, and also increase the proportion of asymptomatic *incidence*. Our findings suggest that neglecting differences in the time profile of transmission between symptomatic and asymptomatic individuals can systematically bias estimates of disease severity, not only during the initial growth phase [25] but also over the course of the epidemic.

We extended the model framework to include the effects of age-dependent heterogeneity in disease severity and assortativity in mixing patterns. When the disease is allowed to spread unchecked, the effects of susceptible depletion dominates the dynamics, resulting in an increase in the mean age of infection through time. However, when we account for the possibility that the asymptomatic individual infections may be longer, the proportion of new infections attributable to transmission from younger individuals increases during the decay phase, tending to decrease the mean age of infection; if asymptomatic infections are long and the epidemic is controlled by intervention or behavior change, mean age of infection can be lower in the declining than in the increase phase. Notably, the age distribution of SARS-CoV-2 infections changed in the US and UK during summer 2020 [3, 26]. For example, the median age of cases decreased from a range of 45–50 years of age to a range of 33–37 years from May to June as the number of cases decreased across all four US census regions ([3], Figure S6). Previous studies primarily attributed these changes to behavioral effects; however, our analysis shows that individual-level differences in transmission dynamics of asymptomatic and symptomatic individuals could have also contributed to these changes.

Our study comes with a number of caveats. Throughout, we considered an idealized intervention which reduces transmission rate by a fixed amount, but real interventions will be more complex. Some interventions, such as contact tracing and self-isolation, are more likely to reduce late transmission by symptomatic individuals and therefore lead to bigger differences between symptomatic and asymptomatic individuals. Other interventions, such as frequent mass testing, will have similar effects on symptomatic and asymptomatic individuals by an equal amount but may still have qualitatively different effects from the intervention we considered (which assumes generation interval distributions are not affected by intervention). Nonetheless, major interventions that drove the current pandemic (e.g., social distancing, mask wearing, and vaccination) are expected to be similar to the idealized intervention we considered. As a result, we expect our qualitative result to be broadly applicable in scenarios where asymptomatic individuals transmit for a longer amount of time. Finally, we emphasize that virus-driven correlations (i.e., asymptomatic transmission is more likely to result in asymptomatic infection) are distinct from demographic correlations (i.e., younger individuals are more likely to infect younger individuals due to assortative mixing). We considered these two correlations separately for simplicity, but both correlations may be present in an actual epidemic: that is, young individuals infected by young asymptomatic infectors may be more likely to remain asymptomatic than those infected by young symptomatic infectors. Coupling of both correlations may further amplify changes in the amount of asymptomatic transmission and incidence over the course of an epidemic.

The dynamics of asymptomatic transmission remain uncertain, despite SARS-CoV-2 having spread throughout the world for over 2 years. More work is needed to better characterize the course of asymptomatic infections with respect to both transmission potential and the duration of infection. Accounting for these sources of individual variation along with the effects of mitigation may aid in understanding how the relative contribution of asymptomatic infections shape epidemic dynamics and in improving the development and deployment of effective control measures.

## Acknowledgments

The authors thank Guanlin Li for contributions to early model development incorporating correlations and David Demory for code review.

## Data Availability

The codes and data are available for download at: https://github.com/Jeremy-D-Harris/Asymptomatic_Transmission_COVID

## Supplemental Information

### Supplemental Tables

**Table S1.** Age-dependent Contact Matrix. Contact matrix, *C*_*n,m*_, includes baseline contact rates in Shanghai with individuals grouped by age into ten year intervals. The average number of contacts per day recorded by the survey participant (rows) is stratified by the age group of the reported contact (columns).

|  |  | Age of Contact |  |  |  |  |  |  |  |
| --- | --- | --- | --- | --- | --- | --- | --- | --- | --- |
|  |  | 0-9 | 10-19 | 20-29 | 30-39 | 40-49 | 50-59 | 60-69 | 70+ |
| Age of Participant | 0-9 | 4.18 | 0.47 | 0.35 | 1.06 | 0.18 | 0.28 | 0.60 | 0.74 |
|  | 10-19 | 0.30 | 9.66 | 0.61 | 1.43 | 1.16 | 0.50 | 0.22 | 0.81 |
|  | 20-29 | 0.08 | 0.11 | 2.18 | 2.83 | 2.65 | 1.65 | 0.74 | 1.05 |
|  | 30-39 | 0.31 | 0.22 | 1.61 | 3.50 | 2.67 | 1.62 | 0.87 | 0.90 |
|  | 40-49 | 0.06 | 0.34 | 1.42 | 2.72 | 2.68 | 1.36 | 0.48 | 1.52 |
|  | 50-59 | 0.09 | 0.12 | 1.17 | 1.95 | 1.95 | 1.90 | 1.28 | 2.68 |
|  | 60-69 | 0.04 | 0.08 | 0.49 | 1.03 | 0.88 | 1.30 | 1.78 | 2.46 |
|  | 70+ | 0.08 | 0.06 | 0.20 | 0.41 | 0.72 | 0.78 | 1.48 | 5.56 |

### Supplemental Figures

**Figure S1.**
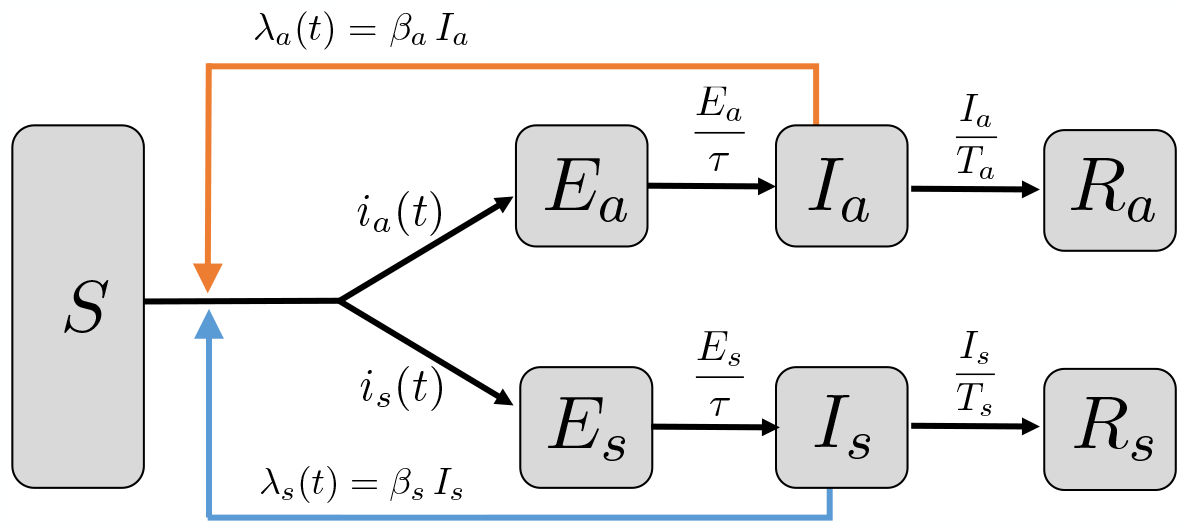
Model structure of SEIR models with asymptomatic and symptomatic transmission. Here, *λ*_*a*_(*t*) is the force of infection due to asymptomatic transmission (orange arrow), whereas *λ*_*s*_(*t*) is the force of infection due to symptomatic transmission (blue arrow). Incident infections remain in an exposed period (latent stage) given by *τ*, which is assumed to be the same for both asymptomatic and symptomatic infections. The asymptomatic (symptomatic) infectious periods, *T*_*a*_ (*T*_*s*_), may differ (Methods). Asymptomatic (symptomatic) incidence is denoted *i*_*a*_(*t*) (*i*_*s*_(*t*)) and differs based on assumptions on correlations between transmission and disease. With fixed proportion of asymptomatic incidence, *p*, transmission and disease are uncorrelated. The asymptomatic incidence is *i*_*a*_(*t*) = *p* (*λ*_*a*_(*t*) + *λ*_*s*_(*t*)) *S*, and the symptomatic incidence is *i*_*s*_(*t*) = (1 *− p*) (*λ*_*a*_(*t*) + *λ*_*s*_(*t*))*S*. With correlations between transmission and disease, the asymptomatic incidence is *i*_*a*_(*t*) = (*p*_*a*|*a*_ *λ*_*a*_(*t*) + *p*_*a*|*s*_ *λ*_*s*_(*t*))*S*, and the symptomatic incidence is *i*_*s*_(*t*) = ((1 *− p*_*a*|*a*_) *λ*_*a*_(*t*) + (1 *− p*_*a*|*s*_) *λ*_*s*_(*t*))*S*. For the age-dependent model, asymptomatic incidence is 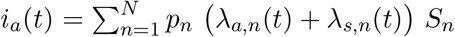 and symptomatic incidence is 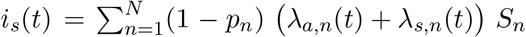, where *p*_*n*_ is the proportion of asymptomatic incidence for age group *n* and the forces of infection are given in Eq. (8).

**Figure S2.**
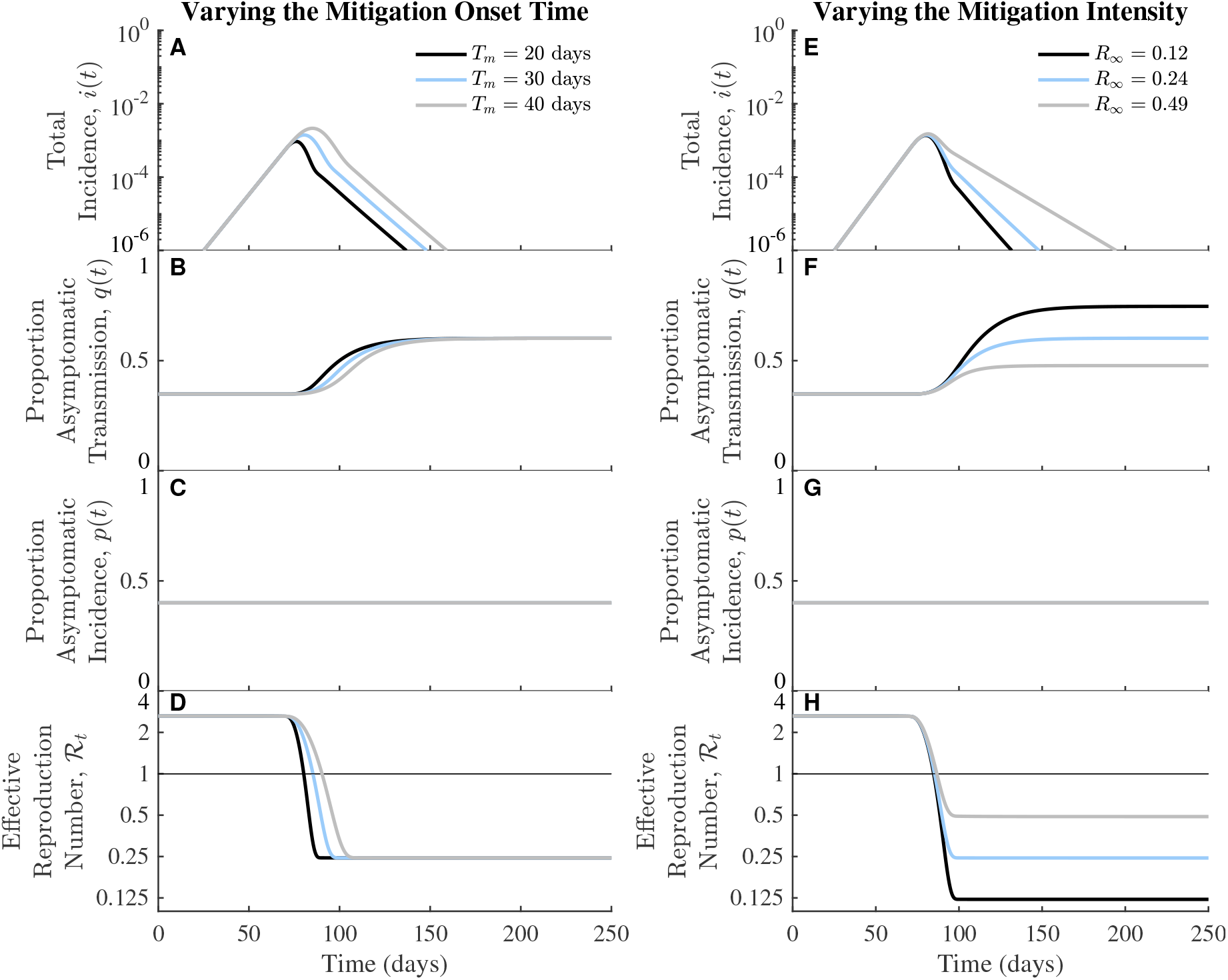
The mitigation intensity determines the magnitude of changes in the realized proportion of asymptomatic transmission over time. For comparison, light blue curves are the same as in Figure 1. (A-D) Fixing the mitigation intensity (i.e., the final effective reproduction number) and varying the mitigation onset time (i.e., duration till full mitigation intensity): 20 days (black), 30 days (light blue), and 40 days (gray). (E-H) Fixing the onset time, while varying the mitigation intensity: *ℛ*_*∞*_ = 0.12 (black), *ℛ*_∞_ = 0.24 (light blue), *ℛ*_∞_ = 0.49 (gray).

**Figure S3.**
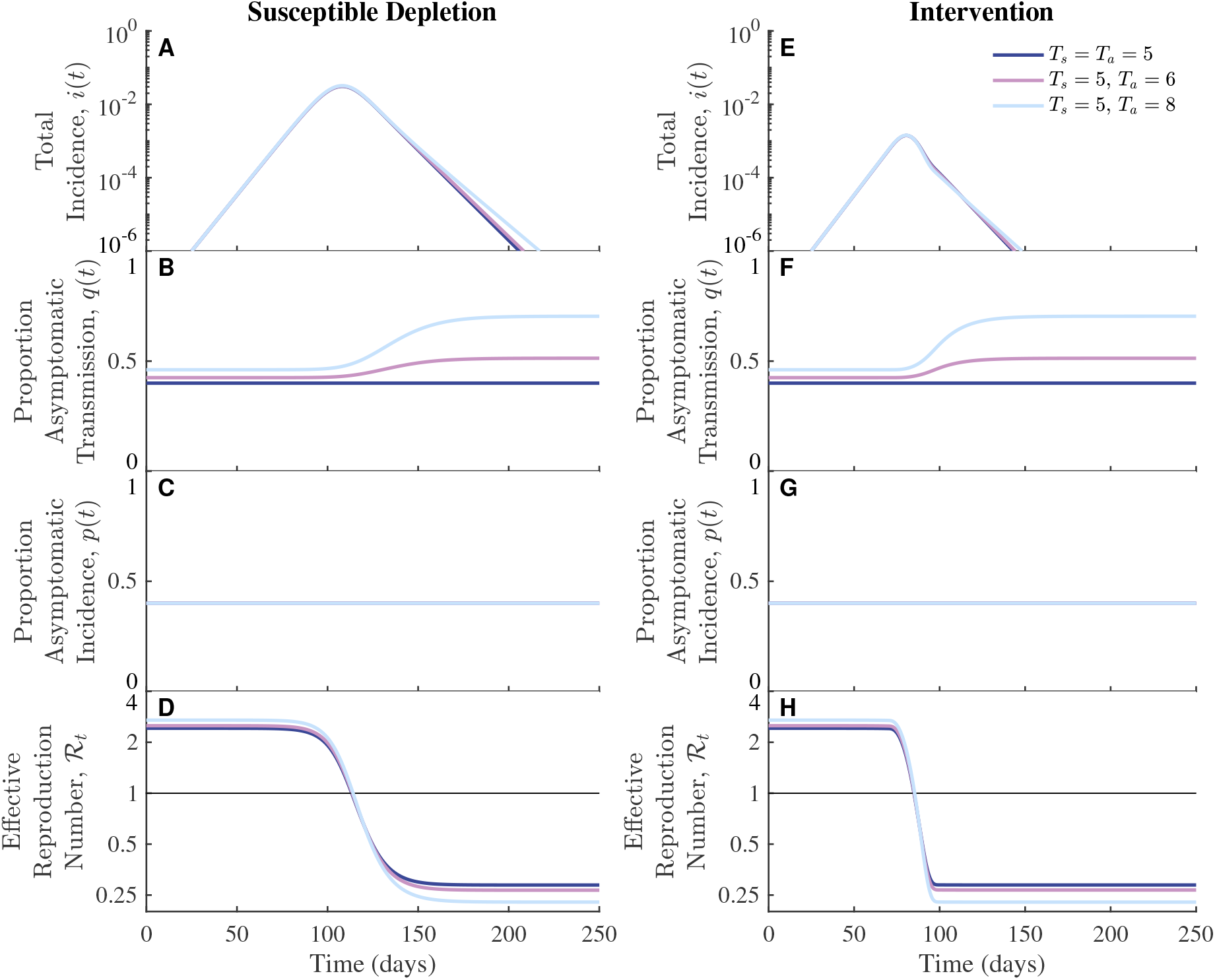
Similar to Figure 1 but assuming asymptomatic and symptomatic infections have the same transmission rates. We fix the symptomatic infectious period, *T*_*s*_ = 5 days and increase the infectious period of asymptomatic infections: *T*_*a*_ = 5 days (dark blue), *T*_*a*_ = 6 days (purple), *T*_*a*_ = 8 days (light blue). The realized proportion of asymptomatic transmission increases over time with similar magnitude as in Figure 1. In contrast to Figure 1, here the initial proportion of asymptomatic transmission is less than the intrinsic proportion, *p* = 0.4, when the generation intervals of asymptomatic transmission are longer than those of symptomatic transmission. Across all simulations, the intrinsic proportion of asymptomatic incidence is *p* = 0.4, and the exponential growth rate is *r* = 0.14 days^*−*1^ (Methods). Other parameter values: *ℛ*_0_ = 2.42, *β*_*a*_ = *β*_*s*_ = 0.484 days^*−*1^ (dark blue); *ℛ*_0_ = 2.51, *β*_*a*_ = *β*_*s*_ = 0.464 days^*−*1^ (purple); *ℛ*_0_ = 2.70, *β*_*a*_ = *β*_*s*_ = 0.436 days^*−*1^ (light blue).

**Figure S4.**
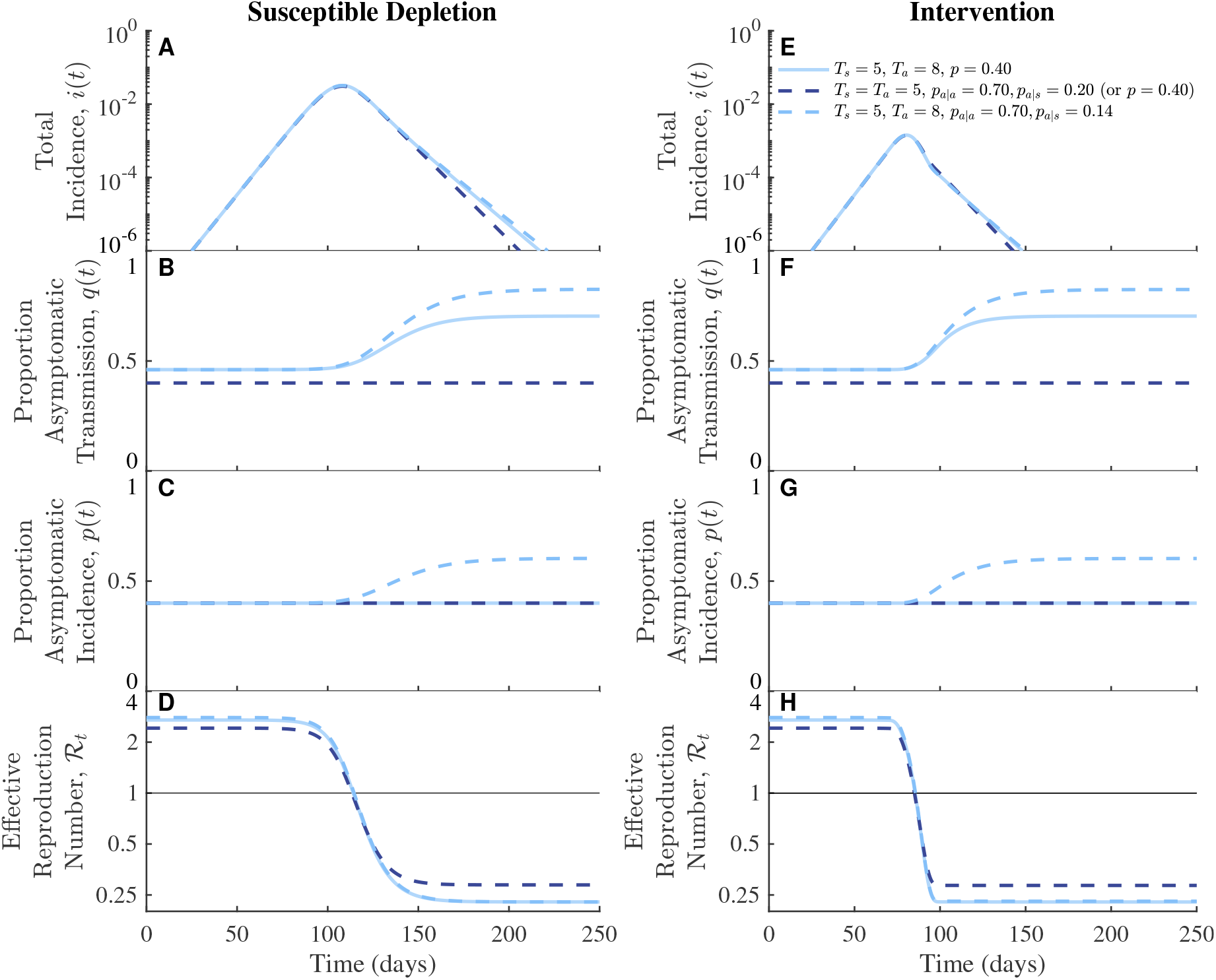
Similar to Figure 2 but assuming asymptomatic and symptomatic infections have the same transmission rates. We fix *T*_*s*_ = 5 days and increase the infectious period of asymptomatic infections from *T*_*a*_ = 5 days (dark blue) to *T*_*a*_ = 8 days (light blue). For comparison, we include simulations with fixed intrinsic proportion of asymptomatic transmission, *p* = 0.4, when *T*_*a*_ = 8 days (solid light blue, the same as in Figure S3). When generation intervals are equal, correlations between transmission and disease status can be included (*T*_*a*_ = 5 days, dashed dark blue) such that the dynamics are identical to those in the case with fixed proportion, i.e., with ‘*p* = 0.4’. When generation intervals differ, correlations cause changes in the realized proportions of asymptomatic transmission and incidence with magnitudes similar to those in Figure 2. Parameter values: *p* = 0.40 (solid light blue), *p*_*a*|*a*_ = 0.70, *p*_*a*|*s*_ = 0.20 (dashed dark blue); *ℛ*_0_ = 2.79, *p*_*a*|*a*_ = 0.70, *p*_*a*|*s*_ = 0.14 (dashed light blue). Other parameter values are the same as in Figure S3 for corresponding colors.

**Figure S5.**
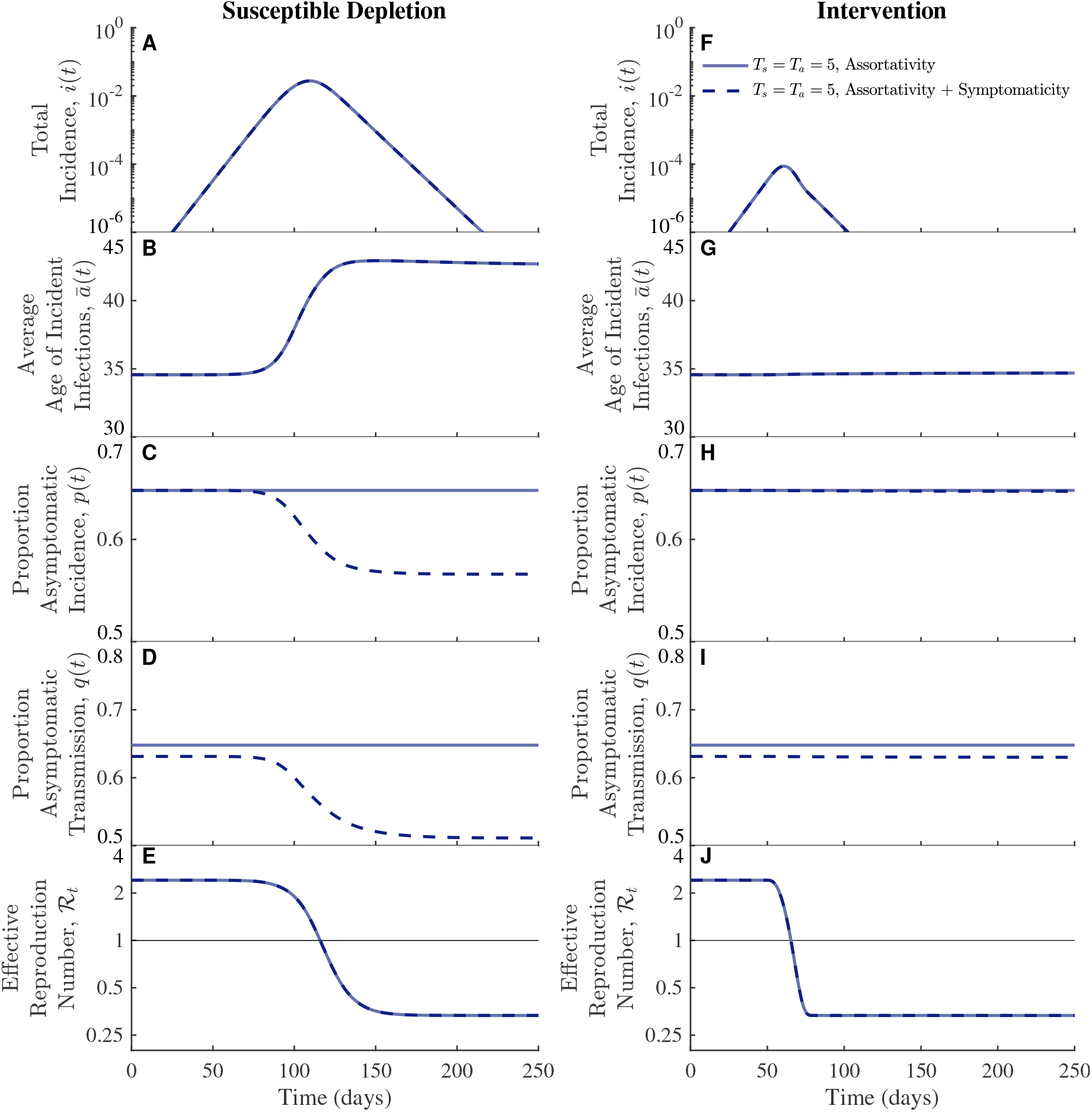
The effects of introducing varying proportions of asymptomatic incidence by age. (A-E) Susceptible depletion. (F-J) Intervention. Coupling age-dependent assortativity with variation in symptomaticity causes decreases in the realized proportions of asymptomatic transmission (C,H) and incidence (D,I). These changes are diminished with intervention (I). Across all simulations, the intrinsic proportion of asymptomatic incidence is 0.648, and the exponential growth rate is *r* = 0.14 days^*−*1^ (Methods). Other parameter values: *ℛ*_0_ = 2.41, *β*_*a*_ = *β*_*s*_ = 0.0445 days^*−*1^.

**Figure S6.**
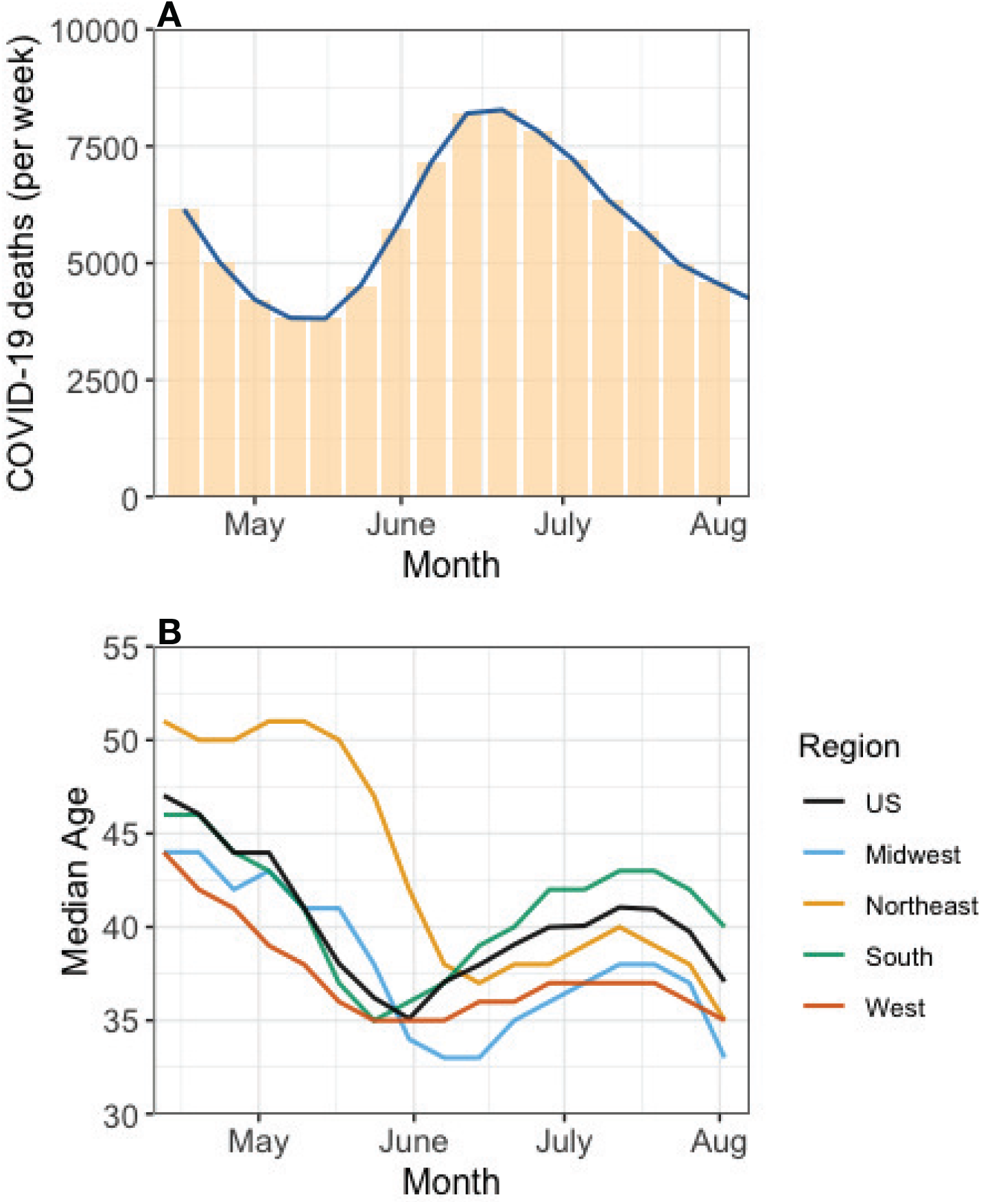
Changes in Median age of COVID-19 infections in US (May-Aug. 2020) as epidemic burden changes over time. **(A)** End of week deaths due to COVID-19 (bars) from 24 May 2020 to 19 September 2020 (CDC surveillance data). Data are shifted by 21 days to estimate the shape of incident infections from May-August 2020. **(B)** Positive RT-PCR tests reported to the CDC by median age from 3 May 2020 to 23 August 2020 from overall US and four US census regions (Data reproduced from MMWR, October 2020 [3]). Tick marks with monthly labels correspond to the 22^*nd*^ of each month.

